# Simulating SARS-CoV-2 epidemics by region-specific variables and modeling contact tracing app containment

**DOI:** 10.1101/2020.05.14.20101675

**Authors:** Alberto Ferrari, Enrico Santus, Davide Cirillo, Miguel Ponce-de-Leon, Nicola Marino, Maria Teresa Ferretti, Antonella Santuccione Chadha, Nikolaos Mavridis, Alfonso Valencia

**Affiliations:** FROM Research Foundation, Papa Giovanni XXIII Hospital, Bergamo, Italy; Bayer, Decision Science & Advanced Analytics for MA, PV & RA Division; Barcelona Supercomputing Center (BSC), C/ Jordi Girona 29, 08034, Barcelona, Spain; Women’s Brain Project (WBP), Gunterhausen, Switzerland; Università di Foggia, Dipartimento di Scienze Madiche e Chirurgiche; Interactive Robots and Media Laboratory (IRLM), United Arab Emirates; ICREA, Pg. Lluís Companys 23, 08010, Barcelona, Spain

## Abstract

Targeted contact-tracing through mobile phone apps has been proposed as an instrument to help contain the spread of COVID-19 and manage the lifting of nation-wide lockdowns currently in place in USA and Europe. However, there is an ongoing debate on its potential efficacy, especially in the light of region-specific demographics.

We built an expanded *SIR* model of COVID-19 epidemics that accounts for region-specific population densities, and we used it to test the impact of a contact-tracing app in a number of scenarios. Using demographic and mobility data from Italy and Spain, we used the model to simulate scenarios that vary in baseline contact rates, population densities and fraction of app users in the population.

Our results show that, in support of efficient isolation of symptomatic cases, app-mediated contact-tracing can successfully mitigate the epidemic even with a relatively small fraction of users, and even suppress altogether with a larger fraction of users. However, when regional differences in population density are taken into consideration, the epidemic can be significantly harder to contain in higher density areas, highlighting potential limitations of this intervention in specific contexts.

This work corroborates previous results in favor of app-mediated contact-tracing as mitigation measure for COVID-19, and draws attention on the importance of region-specific demographic and mobility factors to achieve maximum efficacy in containment policies.

## 1 Introduction

Severe Acute Respiratory Syndrome coronavirus 2, or SARS-CoV-2, is a novel coronavirus strain, discovered in 2019, responsible of a severe respiratory illness named COVID-19 (Coronavirus Disease 2019) that has been declared a public health emergency since January 2020. As of August 18th 2020 more than 13 millions people worldwide have developed COVID-19 and more than 773,000 have died from it.

The spread of COVID-19 has raised new challenges for healthcare systems all over the world, hitting with particular strength Europe and USA, after China. According to the available data, Italy had among the highest number of contagions and dead toll from COVID-19, with over 200,000 confirmed cases and more that 30,000 deceased as of May 14th 2020. However, the spread of COVID-19 has been quite heterogeneous in speed, reach, and lethality, not only from country to country, but also in different regions of the same country. The main possible explanation is given by the delay between the onset of the epidemic, the first diagnosis and the kick-off of containment measures. Other reasons may be due to region-specific variables, such as population density and mean age, societal structure and behaviors. A third factor depends also on the adopted policies for containment and testing, in particular for what concerns the fraction of infectious individuals that never display symptoms (asymptomatic).

In fact, it is now known that the course of infection includes an incubation pre-symptomatic period, during which the patients shows no sign of disease but are still potentially infectious (likely to a lesser degree), after which some individuals progress to a symptomatic state, while others remain asymptomatic, but still potentially infectious[8]. This behavior has important consequence on how we model COVID-19’s epidemiology and plan countermeasures.

Most countries dealing with the epidemics have resorted to nation-wide lock-downs and social distancing to slow down the outbreak; the strategy has been effective in slowing down the epidemic, but lock-downs are temporary measures by nature and simply releasing them without any other parallel containment strategy could very well lead to a new increase of cases. Dedicated works in the scientific literature show that this scenario is avoidable as long as extensive policies of testing and contact tracing are adopted[4, 19].

However, the effectiveness of such measures in controlling the epidemic strictly depends on efficiency in isolating positive and symptomatic cases, as well as their contacts. Earlier work on the topic showed that efficient outbreak control may require tracing and isolation of up to 80% of the contacts, and with very short delay from onset to isolation[6]. This is hardly feasible on a large scale using only traditional methods for contact tracing, and managing the epidemics in the long term will likely require the use of information technology to help implement measures of containment and mitigation. In particular, precise identification of cases and contact tracing and isolation can hardly be performed with traditional methods, and the use of targeted phone apps could highly improve the efficiency of these processes, as shown by the experience of multiple Asian countries - such as South Korea.

Different infrastructures and working interfaces for such an instrument have already been proposed[1], and its potential impact on the virus’s reproductive rate has also been studied[3, 15, 14]. Tracing apps could play a key role in ensuring that the epidemic remains sustainable on the healthcare systems, not exceeding their capabilities, which would otherwise lead to excess mortality.

South Korea pioneered in this approach, both by suggesting to the public the use of applications able to notify contacts with infected subjects, and by mandatory installation, for travellers, of an app implemented by the Ministry of Health and Welfare to monitor COVID-19 symptoms and quarantine for 14 days. Another app, by the Ministry of Interior and Safety, was aimed at monitoring trajectories and contacts of infected subjects and to apply self-surveillance and self-quarantine[11].

Despite concerns over potential privacy violations, the south korean approach managed to achieve efficient control of the epidemics in its earliest phases, and represents a good example of how such an approach could work.

Notable efforts have been done by epidemiologist to model epidemic trends for COVID-19 and the effect of containment measures.

In this proof-of-concept study we built a comprehensive framework to model the COVID-19 epidemic, taking into account population density, the different contributions of symptomatic, pre-symptomatic and asymptomatic contagions, and using it to test the efficacy of targeted interventions such as the aforementioned contact tracing app. In contrast to previous work [15, 14] aimed at modeling the two-way dynamic between individual behaviors and containment policies, we chose to build a global compartmental modeling framework that can account for region-specific factors, such as the effect of population density on contact rate or the role of expected compliance to containment procedures.

Our research builds a model that allows testing the effect of both case isolation and app-mediated interventions in a region-specific fashion.

## 2 Materials and Methods

We built an improved Susceptible-Infectious-Recovered (*SIR*) model[7] with the aims of a) faithfully reproducing the dynamics of the SARS-CoV-2 epidemics, including the respective roles of asymptomatic infection and population density; b) testing the effects of distinct interventions, and specifically the use of phone apps for contact tracing. As Italy was the first western country affected by the SARS-CoV-2 and for which region-specific and intervention-related data was readily available, our analysis is focused on the Italian case. This has allowed us to study how the virus spreads at a very different pace in different Italian districts.

The typical *SIR* model assumes that some degree of immunity, at least temporary, is acquired after SARS-CoV-2 infection; therefore, it is assumed that subjects move from the *S* (Susceptible) compartment to the *I* (Infectious) compartment, and from there, with a daily rate equal to the invers of recovery time, to the *R* (Recovered) compartment, until the relative densities of *S* and *I* become too low for the epidemic to continue. In order to simulate the behavior of asymptomatic and pre-symptomatic individuals, an *A* (Asymptomatic) and *P* (Pre-symptomatic) compartment were added to the model.

To simulate the effect of targeted quarantine measures, we introduced a series of *Q*^∗^ compartments indicating the number of subjects that are quarantined and their status regarding the disease. To model the reversible transition from the different compartment into the state specific quarantine compartments, there are four different *Q* compartments: *QS*, *QI*, *QA* and *QP*. Whenever they are infected, individuals are supposed to move from the *S* compartment to the A or P compartments, with respective probabilities *p_a_* and *p_i_* =1 − *p_a_*. Subjects in the *P* compartments then transfer to the *I* compartment with a daily rate of 1*/τ_i_*, where *τ_i_* is the incubation period. We also assume that asymptomatic and pre-symptomatic patients are less infectious than symptomatic individuals by a factor *f*.

However, there is evidence that pre-sympromatic individuals are highly infectious already about *τ_d_* = 2 days before symptom onset [5], and the focus of our model is transmission dynamics, rather than symptoms manifestations. Under this profile, a *P* subject is already akin to a *I* subject *τ_d_* days before symptom onset. Therefore, we let *P* subjects move into *I* compartment after an incubation period that is *τ_d_* = 2 days shorter, while at the same time increasing the recovery period by the same amount.

Our model assumes the use of a phone app that keeps track of contacts and, once a symptomatic case is identified, notifies the event to everyone who had contacts with them in the pre-symptomatic period, so that they can enter a voluntary quarantine (Figure 1). Such an application is heavily reliant, on one hand, on effectiveness of case isolation on the part of the authorities, and on the other hand on compliance and widespread use of the system by the population. Thus, we assume, for our model, that a *J* fraction of symptomatic cases is identified and undergoes perfect quarantine with zero contacts. We call *j* the fraction of the population using the app and we assume that once in quarantine, they reduce the contact rate by a factor *q*. Therefore, *j*^2^ is the fraction of contacts that happen between individuals using the app. Quarantined subjects that develop the infection undergo the entire course of the disease, whereas those that were not infected in the contact eventually exit quarantine after the quarantine period *τ_q_* (15 days) and become Susceptible again. In the classic SIR model, the rate of transfer between the *S* and *I* compartments depends on transmission rate *β*, that is, the product of number of contacts per subject, *C*, and probability of transmission during a single contact *µ* [13].

**Figure 1:**
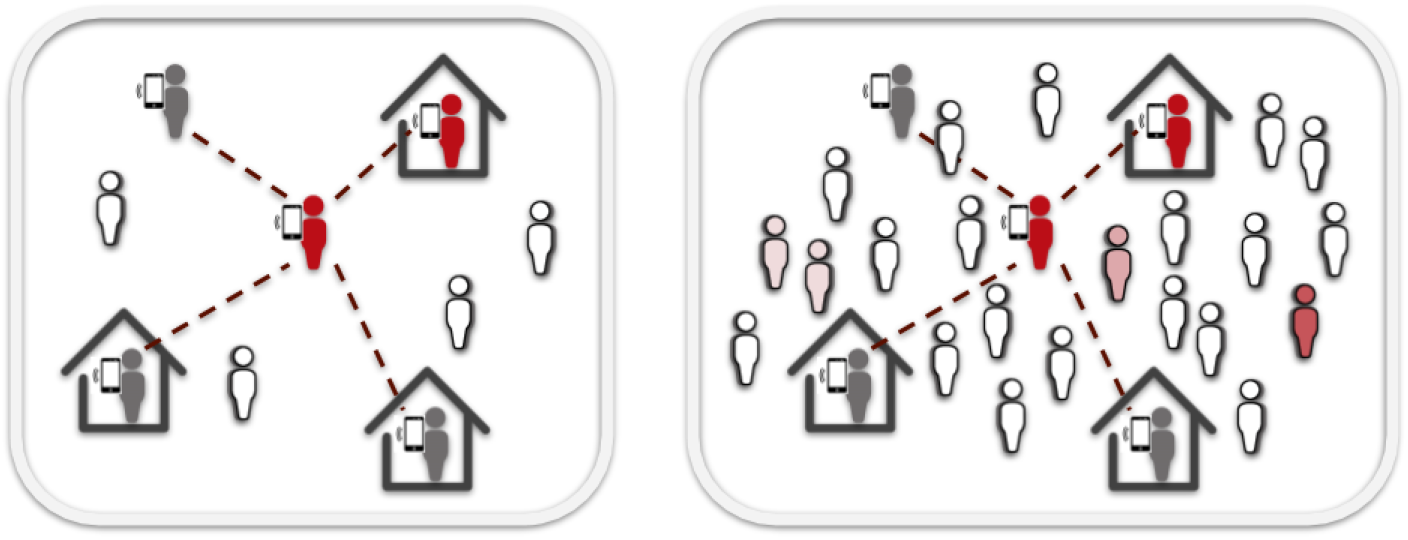
Density and mobility influence contact tracing app effectiveness. Compared to low density regions (left), epidemics may be significantly harder to contain through contact tracing apps in areas with very high population density (right).

To estimate contact rate as a function of population density we built on previous results by Rhodes and Anderson [21] who derived a formula for estimation of daily contact rate of a subject in a population with density *ρ* moving with velocity 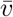 as

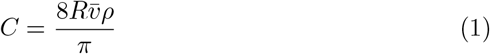

Where *R* is the minimum distance within which two individuals can be said to have a “contact”; for COVID-19, and other air-borne diseases transmitted by droplets expelled from nose or mouth, it is commonly estimated as 1*m* [24].

In our model we assume that Susceptible subjects move to the *P* compartment with a rate proportional to the probability of meeting an *A*, *I* or *P* subject, assuming that at least one of the two does not use the tracing app and/or the case is not successfully identified. This is modeled by making the rate dependent on 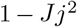. The remaining 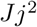 fraction of the contacts between *S* and *I* individuals leads to a transfer from *S* to one of the three *QP*, *QA* and *QS* compartments, each with probability equal to *p_i_*, *p_a_* or 1 − *p_i_* − *p_a_*, i.e. the probability of the contact leading to symptomatic infection, asymptomatic infection or no infection. Individuals in the *QP* compartment eventually transfer to the *QI* compartment after incubation, whereas *QA* and *QS* transfer to *R* and to *S* respectively, with rates equal to 1*/*τ*_i_* and 1*/τ_q_*.

The structure of the model is shown in Figure 2 and it is described by the following set of differential equations. By defining 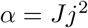 as the proportion of contacts that are successfully contained, we have:

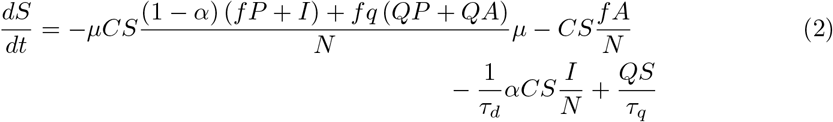

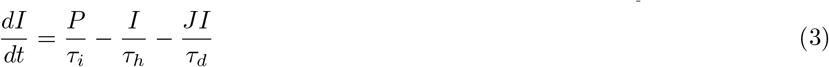

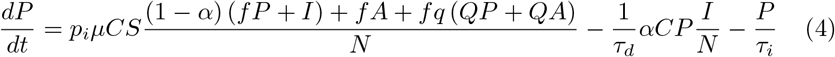

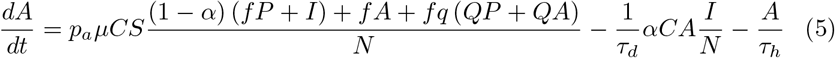

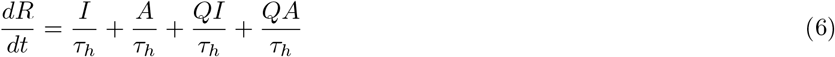

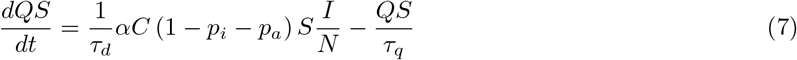

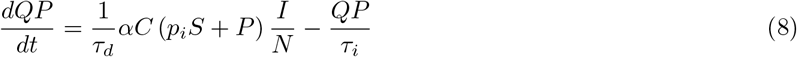

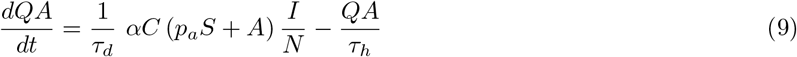

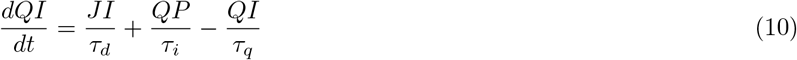

**Figure 2:**
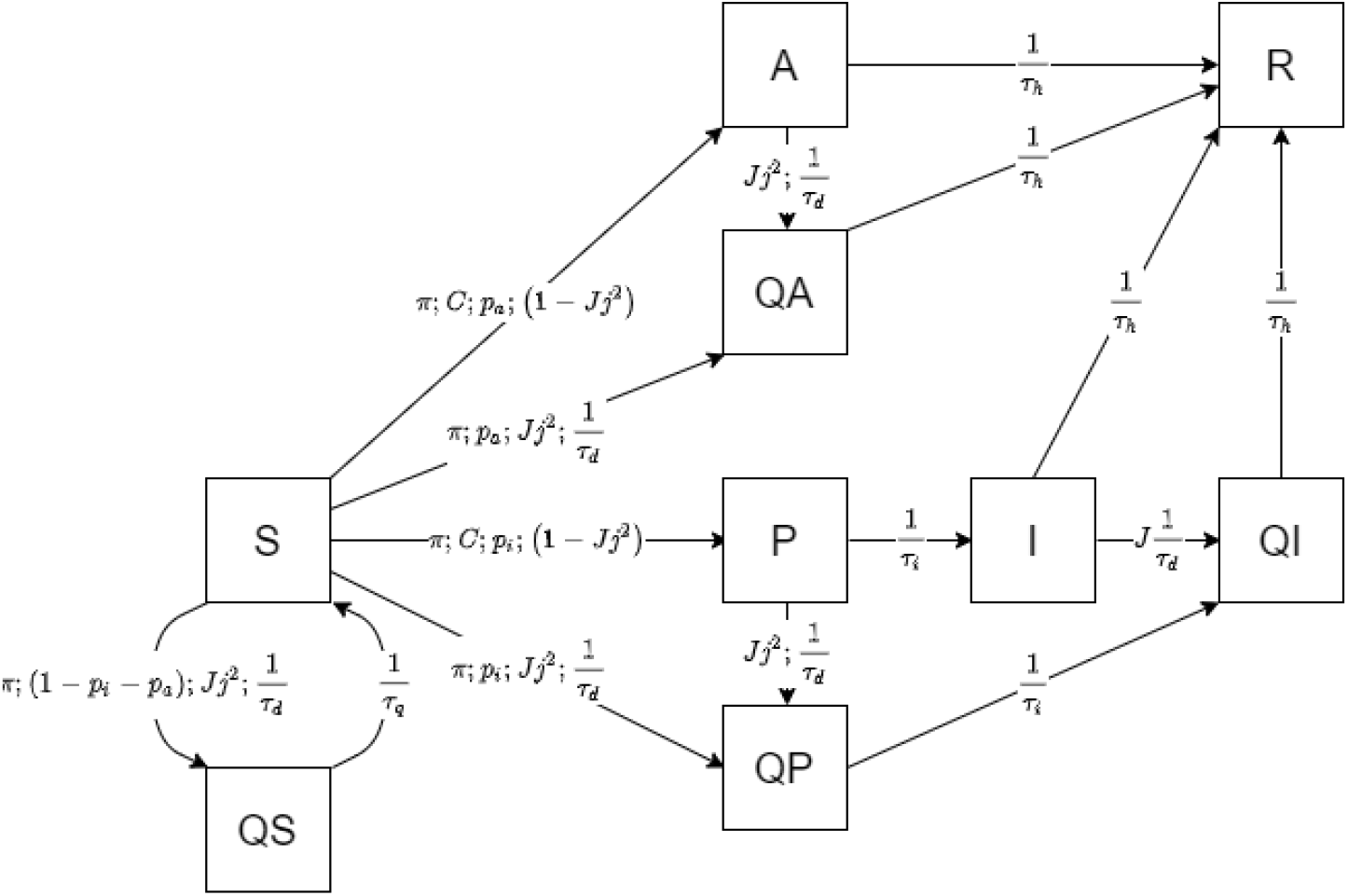
Model outline. Graphical representation of the interactions among the different compartments of the extended SIR model. S, susceptible; I, infected; R, recovered; A, asymptomatic; P, pre-symptomatic; QS, quarantined susceptible; QI, quarantined infected; QA, quarantined asymptomatic; QP, quarantined pre-symptomatic. Rates of transfer between compartments are a function of the parameters annotated on the arrows. A detailed description of such parameters is provided in “Materials and Methods” and Table 1.

The model is governed by a set of ordinary differential equations (2-10) that depend on different parameters. Model parameters are based on values found in different bibliographic references (see Table 1).

**Table 1:** Model parameter values and sources

| Parameter | Description | Value | Source |
| --- | --- | --- | --- |
| $\mu$ | Probability of transmission in a single contact | 0.10 | [2] |
| $p_a$ | Probability of infection being asymptomatic | 0.40 | [3, 10] |
| $f$ | Relative infectiousness of a- and pre-symptomatic compared to symptomatic | 0.10 | [3] |
| $q$ | Contact reduction in quarantine | 0.10 | [18] |
| $\tau_h$ | Time from symptom onset to recovery (days) | 10 | [12] |
| $\tau_i$ | Incubation period (days) | 5.1 | [9] |
| $\tau_q$ | Quarantine duration (days) | 15 | [17] |
| $C$ | Daily contact rate | 14.8 | [22] |
| $R$ | Maximum distance to qualify a “contact” (meters) | 1 | [23] |

For COVID-19 attack rate *µ* we adopted data from the epidemic in Shenzen, where secondary attack rate was estimated between 0.10 and 0.15 [2]. According to Ferretti et al. the most likely estimates for *f* and *p_a_* are 0.1 and 0.4. The 0.4 estimate for the asymptomatic fraction is corroborated by a recent epidemiological study on COVID-19 prevalence in Veneto [10].

In the simulations, we included some basic vital dynamics by adding a *D* (Deceased) compartment (not shown), for which we assumed an overall mortality of 1% in symptomatics. Mortality estimates for COVID-19 are actually still quite uncertain, but Infectious removal by mortality is not expected to significantly affect epidemic trends.

The parameter estimates have a degree of uncertainty and, most importantly, the body of evidence supporting them is continuously growing. Incubation time *τ_i_*, for example has been recently suggested to be higher than our 5.1 days estimate, at 7.76 [20]. Relative infectiousness of *A* and *P* versus *I*, *f*, is another parameter that has strong uncertainty. For this reason, we performed a sensitivity analyses on the two most uncertain parameters (see Supplementary Information).

The simulations were run using R package *SimInf*, a system for stochastic simulation of compartmental models of epidemics [25].

## 3 Experiments

For our experiments we assumed that *J* = 75% of symptomatic cases are identified and perfectly quarantined at symptom onset, i.e., two days after their infectiousness increases. On the other hand, contacts undergoing voluntary self-quarantine are supposed to reduce their contact rate ten-fold (*q* =0.1).

We simulated 12 scenarios with varying contact rate *C* and, most importantly, assuming a different proportion *j* of app users in the population (0, 0.25, 0.5 and 0.75). Each simulation was run on 110 nodes representing Italian districts (with data on area and resident population updated to 2016), and was repeated 50 times, with globally 5,500 simulations per scenario.

For a first set of simulations we assumed a constant population density for all the nodes; this equals the assumption of a unique transmission rate and, therefore, a unique *R*_0_ for the entire set, so that the only region-specific factor affecting the outcome was relative population. Transmissibility has been previously estimated, based on data from the Diamond Princess outbreak, at 1.48 [22]; accordingly, assuming 0.10 attack rate, in this scenario we set a contact rate equal to 14.8. However, this was calculated in a particular scenario where contact rate was supposedly higher than in normal conditions, thus we simulated two more scenarios with lower contact rates, 10 and 7.5.

A second set of simulations accounted for different population densities across the 110 districts. Here, contact rate was allowed to vary following population density under an assumed average daily distance traveled by the subjects, according to Eq.1. Mobility data from two sample european cities (Berga and Barcelona) show an average daily distance traveled per person varying between 1.2 and 5.2 km (personal communication); starting from these figures we simulated scenarios in which subjects travel, on average, 1.5, 2.5 and 5 km per day.

An interactive R Shiny web application, enabling the exploration of simulation scenarios, is available at https://flowmaps.life.bsc.es/shiny/ct_app/.

## 4 Data

Our simulations were based on population and surface area data from 110 italian districts updated to 2016. This kind of data is subject to frequent administrative rearrangements, but actual demographics have remained substantially stable for the aims of this work, and since some districts have since be grouped together, older data has actually higher spatial resolution.

Table 2 summarizes population data about the five most and least densely populated districts. Over the entire national territory, density was 2.01*/km*^2^, and in districts it ranged from 0.03 to 2.65. Corresponding surface area ranged from 212 to 7400*km*^2^. Globally, population was 60,589,085. Ogliastra was both the least populated and least densely populated district, with 57,185 inhabitants and a density of 0.03*/km*^2^; the district with most residents was Rome (4,353,738 residents), but the most densely populated was Neaples.

**Table 2:** Summary statistic of the five most and least densely populated districts.

| Districts's data summary |  |  |  |  |
| --- | --- | --- | --- | --- |
| District | Region | Density | Surface ($km^2$) | Residents |
| Napoli | Campania | 2.65 | 1171 | 3107006 |
| Monza and Brianza | Lombardia | 2.15 | 405 | 868859 |
| Milano | Lombardia | 2.04 | 1579 | 3218201 |
| Trieste | Friuli-Venezia Giulia | 1.11 | 212 | 234682 |
| Roma | Lazio | 0.81 | 5352 | 4353738 |
| ... | ... | ... | ... | ... |
| Grosseto | Toscana | 0.05 | 4504 | 223045 |
| Olbia-Tempio | Sardegna | 0.05 | 3399 | 160672 |
| Nuoro | Sardegna | 0.04 | 3934 | 156096 |
| Aosta | Valle d'Aosta | 0.04 | 3263 | 126883 |
| Ogliastra | Sardegna | 0.03 | 1854 | 57185 |

## 5 Results

Results of the simulations are summarized in Figures 3 and 4, showing the time curves of the sum of the *I* and *QI* compartments and expected mortality. Clearly the epidemic peak is expected to vary with increasing contact rate, assuming that transmissibility and recovery rate are constant. As expected, in all simulated scenarios, app-aided contact tracing significantly decreased the effective reproductive number *R_t_* and height of the epidemic peak. In our sensitivity analyses, using the slightly longer incubation time had no significant impact in our simulations, whereas the increased infectiousness causes an expected delay in the complete suppression of infections, in particular under conditions of normal mobility.

**Figure 3:**
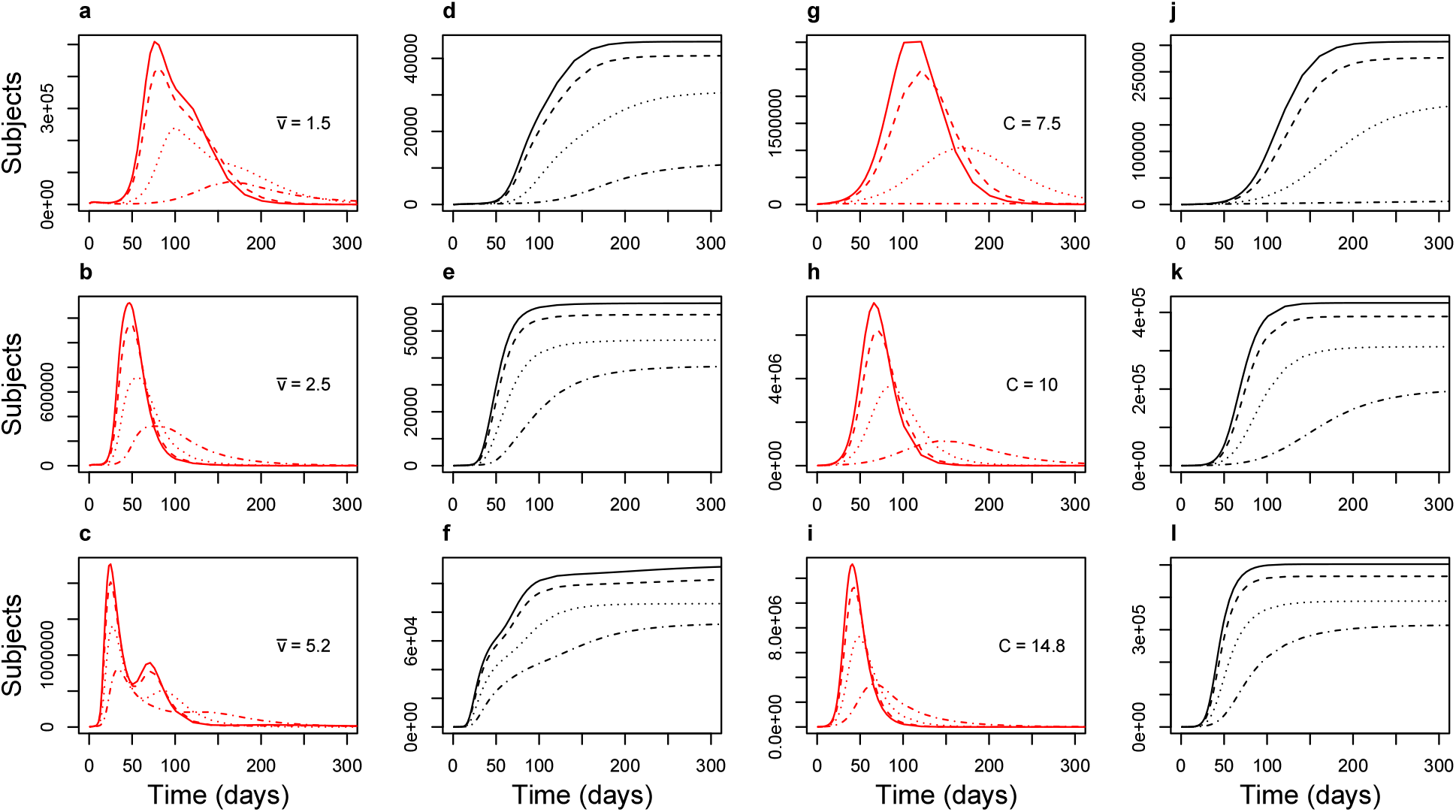
Total symptomatic population (*I* + *QI* compartments) in red and simulated mortality (black) in the 48 scenarios.Density-dependent contact rate simulations on the left (plots a-f);fixed contact rate simulations on the right (plots g-l). Each curve results from the sum over 110 districts averaged over 50 replicates per district. Solid lines represent no app users; dashed, dotted and dashed-dotted lines show increasing fractions of the population using the app (25, 50, 75%). All simulations can be interactively explored at https://flowmaps.life.bsc.es/shiny/ct_app/.

**Figure 4:**
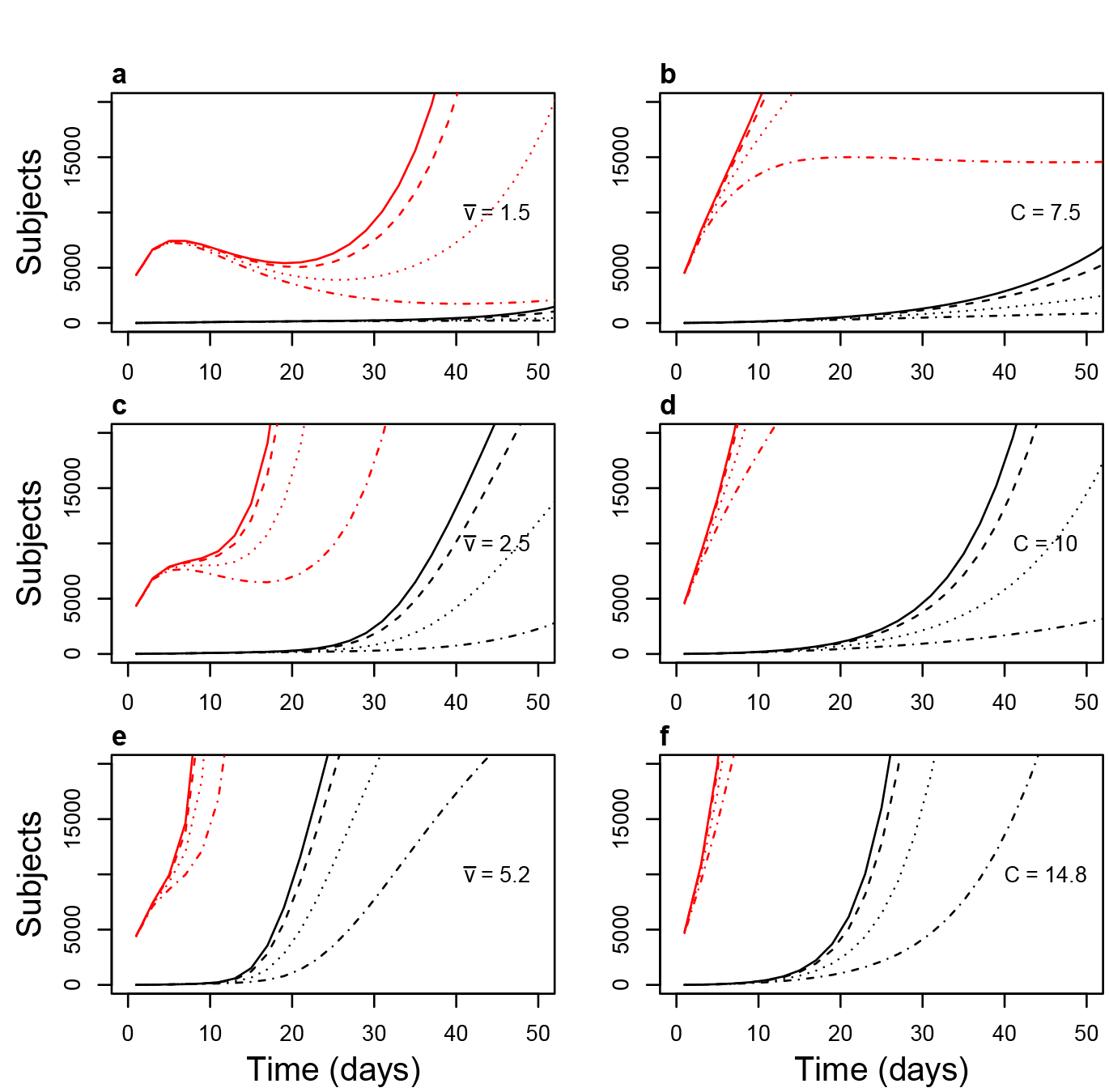
Detail of Figure 3 showing successful/unsuccessful suppression in the first 50 days. Suppression is achieved in the most optimistic scenario (contact rate 7.5, app users 75%) in the scenarios with fixed contact rate (right panels, upper), whereas its success varies from district to district in scenarios with density-dependent contact rate (left panels).

### 5.1 Constant contact rate

In scenarios with constant contact rate equal to 7.5, symptomatic cases isolation was per se very effective in slowing the epidemic, so much so that app-mediated contact tracing managed to achieve suppression even with only 75% (Fig 4 b) of the population using the app and complying to self-quarantine, whereas with 50% peak height was reduced more than 2-fold (Fig 3 b). This shows that, in scenarios with lower baseline contact rate and efficient isolation of cases, app-mediated contact tracing can achieve epidemic suppression.

On the other hand, nation-wide suppression was not achieved in the less optimistic scenarios with 10 and 14.8 contact rate; however, the app induced a very effective mitigation, with peak number of infectious reduced roughly 4-fold in the worst-case scenario and with 75% of the population using the app (Fig. 3 d and f).

### 5.2 Density-dependent contact rate

In scenarios where contact rate was allowed to vary with population density the epidemic trend was, as expected, significantly different from region to region. In Figure 3, left panels, the curves of symptomatic infections over time are shown; compared to simulations with constant contact rate, it is evident the presence of two more or less distinct epidemic peaks; this may reflect the presence of groups of districts with different population densities. However, the distinction tends to disappear with increasing proportions of app users. The simulations show that suppression can be achieved in most regions even with only 25% of the population using the app (dashed lines), but the epidemics does not die out, being almost entirely sustained by the districts with the highest population density (Milan, Monza, Neaples; Fig. 5). This result is, clearly, achieved by using the app to augment an efficient tracking and isolation of new symptomatic cases, and indicates that, as for all interventions, effectiveness of app-mediated contact-tracing and voluntary quarantine should be evaluated in the light of region-specific differences.

**Figure 5:**
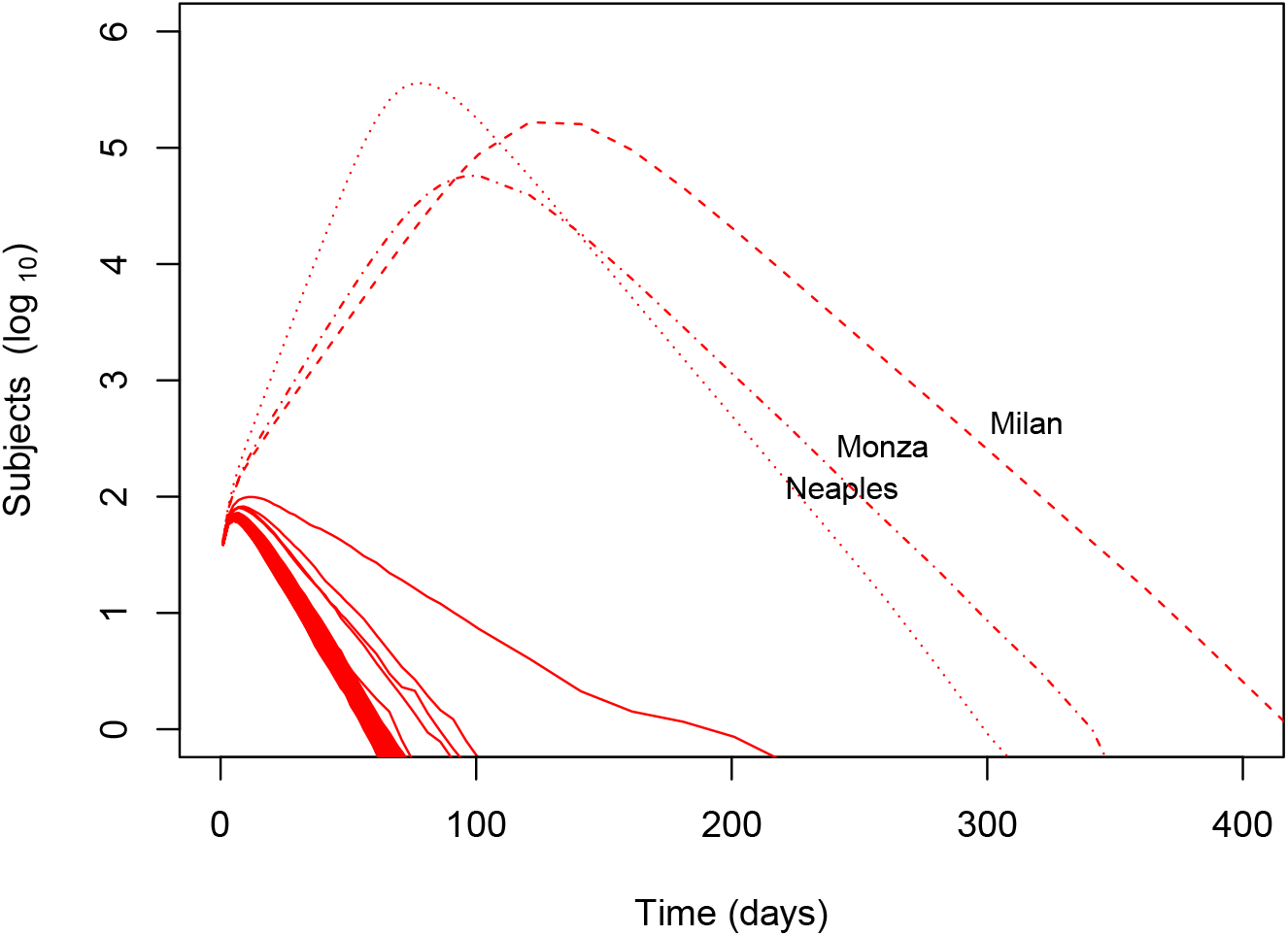
Epidemic curve for individual nodes (districts) in the scenario with average velocity 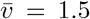 and fraction of app users *j* = 25%. The epidemic outbreak is entirely sustained by the three highest-density districts, whereas in the others the effective reproductive number is below 1.

### 5.3 Quarantine measures impact

The model allows to also keep track of the number of *I* subjects successfully quarantined (“true positives”) and of the subjects that underwent quarantine without actually being infected (“false positives”), by tracing the population in the *QI* (Quarantined-Infectious, subjects that were rightfully quarantined) and *QS* (Quarantined-Susceptible, subjects that were quarantined but did not contract infection) compartments. The maximum number of subjects in each compartment at the same time is summarized in Table 3.

**Table 3:** Average maximum True Positives (*QI*) and False Positives (*QS*) in each scenario.

| | $j$ | $QI$ | $QS$ |
| --- | --- | --- | --- |
| $\bar{v} = 1.5$ | 0 | 411850.46 | 0 |
| $\bar{v} = 1.5$ | 0.25 | 345302.96 | 126772.1 |
| $\bar{v} = 1.5$ | 0.5 | 195800.12 | 304706.56 |
| $\bar{v} = 1.5$ | 0.75 | 58791.4 | 227277.26 |
| $\bar{v} = 2.5$ | 0 | 1082325.62 | 0 |
| $\bar{v} = 2.5$ | 0.25 | 937295.72 | 344843.98 |
| $\bar{v} = 2.5$ | 0.5 | 617449.32 | 958998.18 |
| $\bar{v} = 2.5$ | 0.75 | 282200.52 | 1096092.2 |
| $\bar{v} = 5.2$ | 0 | 1774628.08 | 0 |
| $\bar{v} = 5.2$ | 0.25 | 1641431.7 | 589880.54 |
| $\bar{v} = 5.2$ | 0.5 | 1202505.64 | 1801323.82 |
| $\bar{v} = 5.2$ | 0.75 | 702748.88 | 2656114.82 |
| $C = 7.5$ | 0 | 2482094.86 | 0 |
| $C = 7.5$ | 0.25 | 2029409.8 | 747889.48 |
| $C = 7.5$ | 0.5 | 866790.1 | 1346998.68 |
| $C = 7.5$ | 0.75 | 13639.94 | 51768.96 |
| $C = 10$ | 0 | 5993749.92 | 0 |
| $C = 10$ | 0.25 | 5110050.2 | 1879426.1 |
| $C = 10$ | 0.5 | 3023884.64 | 4691037.66 |
| $C = 10$ | 0.75 | 960177.86 | 3703523.2 |
| $C = 14.8$ | 0 | 10492705.48 | 0 |
| $C = 14.8$ | 0.25 | 9162768.02 | 3395616.34 |
| $C = 14.8$ | 0.5 | 6105113.84 | 9497105.26 |
| $C = 14.8$ | 0.75 | 2977727.44 | 11519918.38 |

Both false positives and true positives are naturally dependent on the success or failure of epidemics suppression, and will be very low when suppression is achieved. In density-dependent scenarios with 75% of app users the maximum number of Susceptible subjects quarantined at the same time ranged from 200,000 to 2,000,000, whereas in scenarios with fixed density the variation was much higher, mainly due to the fact that with contact rate *C* =7.5 and 75% app users suppression is achieved.

As expected, True Positives decrease as app coverage increases and epidemic spread is impaired, but at the same time the number of subjects that are subjected to voluntary quarantine without reason increases. The two quantities cross over between *j* =0.25 and *j* =0.5, where subjects that are wrongly quarantined surpass those that are actually infectious.

## 6 Discussion

### 6.1 Modeling framework

The use of targeted app for contact tracing has been proposed as a means to control the COVID-19 epidemics when lockdown measures are lifted. It has been shown that, given certain combinations of efficacy in case identifications and compliant use of such an instrument, the approach can contribute to the effective reproductive number *R_t_* of the disease below 1 [3]. Here we aimed to model the effect of app-mediated contact tracing taking into account population density and transportation, at the same time making it possible to monitor the number of patients that are quarantined and their status concerning the infection.

This is a much needed approach if we wish to implement precise and timely specific intervention making the infections and contagion sustainable for health care systems.

The model uses a series of *Q*∗ compartments to model the behavior and status of subjects that are quarantined for symptomatic infection or based on contact tracing. To our knowledge, this is the first model that allows simulation and prediction of the outcomes of the epidemic both accounting for differential population density and quarantine measures. This is particularly important since it allows to visualize the effect of contact-tracing apps along the entire duration of the epidemics, and also because the management of the infection has to take into account the specific characteristics of a given region and implement measures accordingly. In fact, the application of containment policies disregarding region-specific conditions can result in measures which are not needed or too drastic. As a consequence, rather than providing support, such policies might result in a burden for the psychological well-being of people as well as detrimental for the economy[16].

Compartmentalization of Quarantined subjects has a remarkable potential usefulness in studying the socio-economic impact of voluntary self-quarantine suggested by app tracing. Clearly, a larger proportion of app users entails better epidemic control. However, it is expected that the number of false positives in the *QS compartmen*t increases accordingly, with increased losses in terms of well-being and resources.

Despite the many parameters considered in our model, our work still falls into a classic compartmental epidemiologic framework; this is a net advantage in terms of interpretability of results and generalizability. To our knowledge, this is also the first model that allows keeping track of subjects that undergo voluntary quarantine.

### 6.2 Contact tracing effectiveness

According to our model, case isolation is *per se* a very effective containment measure that, as long as cases are identified and isolated with a very high success rate, can achieve suppression of the epidemics in a series of theoretical scenarios. However, coupling case isolation with immediate app-mediated contact tracing produces a remarkable improvement in success of the strategy, achieving in all scenarios a very effective mitigation of contagion and, in some scenarios, full suppression. These results highlight the benefit of introducing contact-tracing as a measure of pandemic prevention and control as well as the positive impact that this would have especially upon critical circumstances.

Feasibility of epidemic control by app-mediated contact tracing has been suggested already by Ferretti et al.; by decomposing contributions to *R*_0_ from symptomatic, pre-symptomatic, asymptomatic and environmental sources, they show that *R_t_ <* 1 can be attained for certain combinations of efficiency in case isolation and compliance in contact tracing. Our model allows to directly simulate such scenarios while at the same time keeping track of the trends in isolated and quarantined cases. This is particularly relevant as it allows to quantify the effect of interventions on specific compartments, e.g. it is possible to trace the number of individuals that are quarantined at a specific time-point, a piece of data that is potentially very helpful in designing cost-effectiveness analyses of containment measures.

### 6.3 Density and traveling

Another important result comes from our simulations with a density-dependent contact rate. In our simulation different Italian districts behaved very differently, and in all scenarios suppression was easily attained in the less densely populated regions, whereas it failed in the others (Fig. 5). This is consistent with the different epidemic trends that have been observed to date in Italian districts; however, it must be pointed out that differences between districts may as well be justified by different approaches in dealing with the epidemics, time to first diagnosed case versus numbers of people already infected in the population and not yet recognized and, most importantly, is influenced by the nation-wide/region-wide lockdown put in place by the central government.

It is also of note that by making contact rate dependent on both density and daily distance traveled, our model takes into account the potential effectiveness of policies aimed at optimizing and regulating transportation, especially in high density regions. According to the model, effective suppression of the epidemics in such areas is strongly dependent on such measures.

### 6.4 Limitations and future research

The main limitation of our work is that there is some uncertainty in the parameters that have to be plugged-in the simulations; anyway, we were able to derive reasonable estimates for all of them, and performed a sensitivity analysis to check that our results are robust to changes in the most uncertain parameters. We adopted some credible figures for the asymptomatic/symptomatic infections ratio and for relative decrease in infectiousness in asymptomatic subjects, as well as for probability of infection per contact; however the greatest uncertainty is precisely in estimation of contact rate, as this is a variable that is influenced by specific environmental and cultural factors, e.g. individual mobility, social interactions, transportation systems, as well as general social distancing measures that have been implemented wherever the epidemics took place. Our first choice estimate for contact rate came from the experience of the Diamond Princess, based on which we estimated it at a very impressive figure of 14.8, a condition that would hardly allow for suppression in a nation-wide context. However, it is evident that the situation in a closed environment favors a higher contact rate, thus this number is likely to be significantly overestimated. This makes the scenario a worst-case one, suggesting that, in real world experience, case isolation and contact tracing may be more effective than predicted.

In scenarios where we modeled contact rate as a function of population density, the most relevant scaling factor is 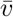, i.e. the theoretical average daily distance traveled by individuals. This is a measure that is not readily estimable, which is why in the present work we showed results for a series of credible scenarios; however, using mobility data from European cities we managed to obtain credible figures to plug-in the model. Our model does take into account interpersonal distance in the form of *R* parameter; a potential expansion of this framework is to account for time spent within the 1*m* interpersonal distance, as well as distinguishing between high-risk (e.g. taxis, buses) and low-risk (e.g. walking, personal car) means of transportation.

However, this work proves the feasibility of including population density and transportation in an expanded *SIR* model, and suggests that, even if travel between districts is forbidden, the epidemics may still be significantly harder to contain in areas with very high population density (for example, in Italy, the districts of Milano, Monza and Napoli). The model can be further improved and expanded by adding age-specific compartments, sex and gender factors, and risk classes, by refining the implementation of vital dynamics, and by modeling different methods for contact tracing with varying degree of compliance.

### 6.5 Conclusions

We approached the model with a simulation based approach; this is computationally intensive but still manageable by most software and hardware and less demanding than stochastic models based on individual data, and allows for a high degree of customizability by fine-tuning its parameters on specific interventions.

The model constitutes a viable framework to monitor epidemic trends and assess the effect of interventions. Our results show that (1) voluntary self-quarantine based on contact-tracing apps, together with efficient case isolation, can give a relevant, and in some scenarios decisive, contribution to epidemics mitigation/suppression; (2) at the same time, the success of this strategy can depend heavily on population density and transportation.

## Data Availability

Data available on request from the authors.

## 7 Acknowledgments

This material is based upon work supported by the European Commission’s Horizon 2020 Program, H2020-ICT-12-2018-2020, “INFORE - Interactive Extreme-Scale Analytics and Forecasting” (ref. 825070).

## 9 Author contributions

A.F., E.S. and N.M. conceived the experiments; A.F. designed the model and ran simulations; A.F. and E.S. wrote the paper; D.C. and M.P-d-L. revised the model and code; D.C. wrote the web app; M.T.S., A.S.C. N.M. and A.V. revised the paper and helped with interpretation and discussion; A.V. supervised model design and provided mobility data. All of the authors approved the final version of the paper.

